# Generalized Prediction of Hemodynamic Shock in Intensive Care Units

**DOI:** 10.1101/2021.01.07.21249121

**Authors:** Aditya Nagori, Pradeep Singh, Sameena Firdos, Vanshika Vats, Arushi Gupta, Harsh Bandhey, Anushtha Kalia, Arjun Sharma, Prakriti Ailavadi, Raghav Awasthi, Wrik Bhadra, Ayushmaan Kaul, Rakesh Lodha, Tavpritesh Sethi

## Abstract

Early prediction of hemodynamic shock in the ICU can save lives. Several studies have leveraged a combination of vitals, lab investigations, and clinical data to construct early warning systems for shock. However, these have a limited potential of generalization to diverse settings due to reliance on non-real-time data. Monitoring data from vitals can provide an early real-time prediction of Hemodynamic shock which can precede the clinical diagnosis to guide early therapy decisions. Generalization across age and geographical context is an unaddressed challenge. In this retrospective observational study, we built real-time shock prediction models generalized across age groups (adult and pediatric), ICU-types, and geographies. We trained, validated, and tested a shock prediction model on the publicly available eICU dataset on 208 ICUs across the United States. Data from 156 hospitals passed the eligibility criteria for cohort building. These were split hospital-wise in a five-fold training-validation-test set. External validation of the model was done on a pediatric ICU in New Delhi and MIMIC-III database with more than 0.23 million and one million patient-hours vitals data, respectively. Our models identified 92% of all the shock events more than 8 hours in advance with AUROC of 86 %(SD= 1.4) and AUPRC of 93% (SD =1.2) on the eICU testing set. An AUROC of 87 % (SD =1.8), AUPRC 92 % (SD=1.6) were obtained in external validation on the MIMIC-III cohort. The New Delhi Pediatric SafeICU data achieved an AUROC of 87 % (SD =4) AUPRC 91% (SD=3), despite being completely different geography and age group. In this first, we demonstrate a generalizable model for predicting shock, and algorithms are publicly available as a pre-configured Docker environment at https://github.com/tavlab-iiitd/ShoQPred.

## Introduction

Shock is one of the most common complications in patients admitted to the ICUs, with an incidence as high as 33%^1^. Hypovolemic, cardiogenic, and septic shock are characterized by altered hemodynamics^2^, presenting as hemodynamic shock (HS). The mortality rate in patients who develop shock in ICU is as high as 34% in developing countries^3^, triggered by a cascade of poor blood perfusion, inadequate oxygen availability to vital organs, and multiple organ failure. Early identification is critical for aggressive management^4,^ improved patient outcomes, and mortality reduction^5,6,7,8^ However, a delay in early identification can compound the risk of mortality and organ failure. These delays can be attributed to a few observations; firstly, there is a plethora of information being generated inside an ICU. For physicians and nurses to observe the data on a real-time basis is overwhelming. Information overload has worsened the physician’s workload, contributed towards burnout, and increased the chances of errors^9^. Studies have found an association between burnout and jeopardized care^10,11,12^. Thus, removing the risk of informational overload using Machine learning based assistive tailored models can prevent the risk of suboptimal care^9^. Patient care also gets compromised as a result of the insufficient strength of nurses or ICU staff. Studies have found that lowering the bed-to-nurse ratio or ICU staffing can result in poor patient outcomes^13,14^. Therefore, the artificial intelligence-based algorithm can help in monitoring and prioritizing deteriorating patients. It is imperative to reduce the information burden by building high-performance decision models that can recognize the characteristic patterns to assist the ICU staff with actionable and interpretable decision-making insights. Hemodynamic instability is one of the initial signs related to the manifestation of further deterioration. Therefore, we built a decision support system that can produce early warning scores with high recall and precision.

Secondly, the development of models for early warning monitoring support systems should have the feasibility of deployment to the same setting and other settings. Although the literature is full of models relying upon information from various sources^15,16,17,18^, their generalization to other settings is a major concern.

In this work, we developed real-time models for early identification of shock using high-resolution vitals time series data using hand-engineered time-series features, machine learning, and deep learning approaches. Vitals time-series are routinely generated at a much higher resolution than hourly nursing notes, hence have the potential to forecast critical outcomes^19 20 21^. Our Safe-ICU data warehouse^22^ with more than 0.23 million hours of patient physiological time-series vitals data allowed us to validate on pediatric age groups.

High-resolution vitals time-series data has shown potential for multicenter generalization^21^, but very few studies have been conducted to evaluate the generalizability of AI models for the ICU^15, 21, 23^. To our knowledge, none of the studies evaluated the potential of models learned on the adult population and generalized to pediatric ones. One of the critical challenges to do so is the dependency on a large set of clinical measurements and differences in age-based criteria. These challenges can be overcome by using high-resolution physiological vitals time-series data generated through monitoring sensors and transfer learning^21, 24^.

In this work, we report machine learning based prediction models, which take readily available time-series vitals data to forecast shock status up to eight hours ahead of its onset. We do this using hand-engineered time-series features derived from the raw signals to build these models (Figure 1). There is a high percentage of patients who developed shock in the ICU. We built cohorts around patients who developed shock after 7 hours of ICU admission. Artificial intelligence algorithms can generalize and scale; therefore, we evaluated our models for their ability to generalize on MIMIC-III adult ICU and SafeICU pediatric ICU data using a transfer learning approach. Thus, our work aimed to utilize readily available ICU time-series data to build robust parsimonious predictive models for the onset of Hemodynamic shock and evaluate the potential for generalization of these models to another setting.

**Figure 1:**
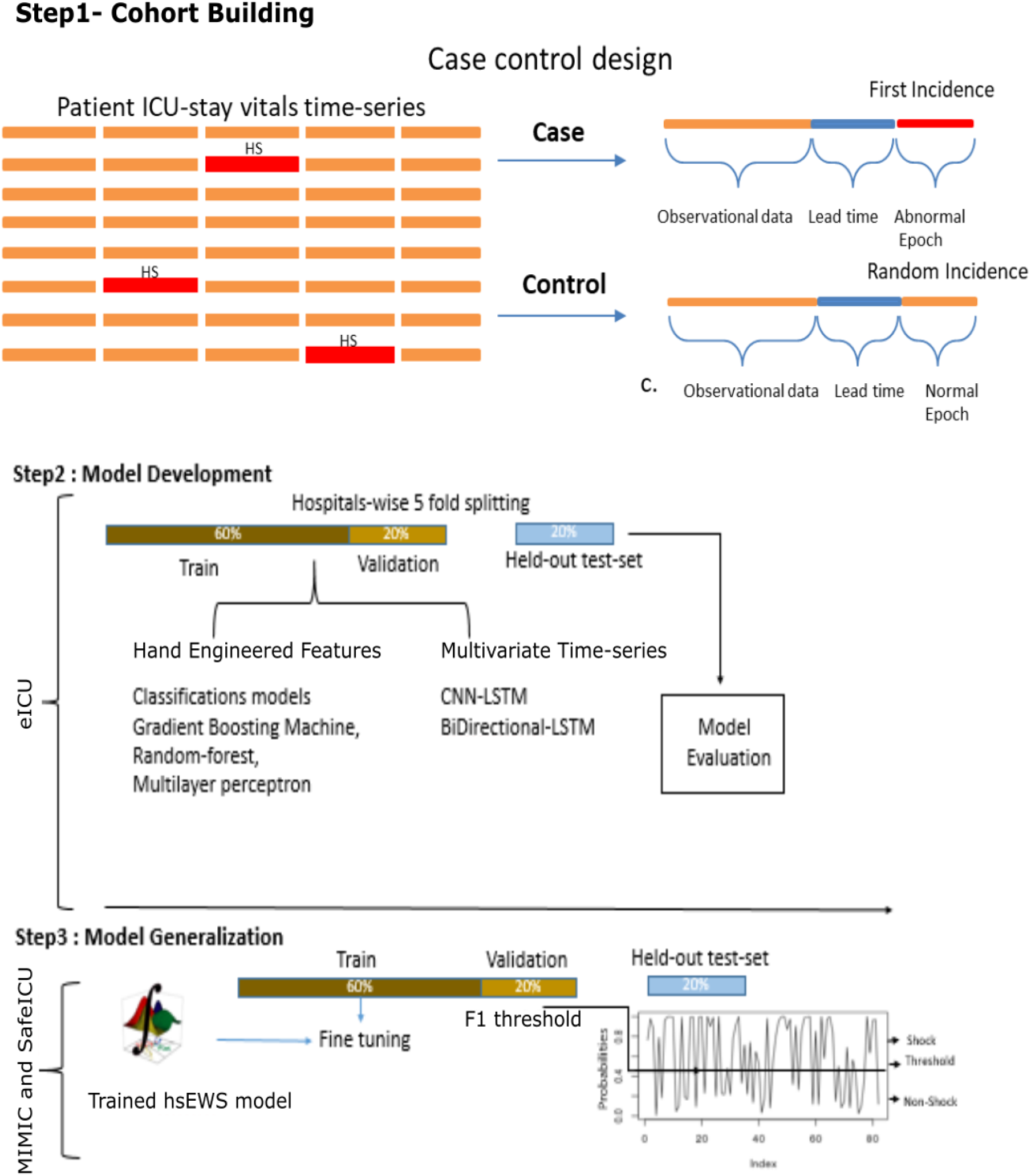
Summary of the pipeline for generalizing the prediction of hemodynamic shock. Step 1 shows Cohort Building. Step 2 shows the model-building to predict shock 0-8 hours in advance and evaluation. Step 3 shows the generalization of learned models on MIMIC-III data and Indian settings’ SafeICU data.

## Results

The periodic vitals data of eICU data was preprocessed according to inclusion and exclusion criteria, resulting in 16,246 admissions (Supplementary figure 1a). The median length of stay of patients in the extracted cohort was 3.1 days. As expected, our data show that mortality was positively correlated with the fraction of length of stay with hemodynamic shock (Pearson’s correlation r = 0.89, p = 4.8*10^−4^) (Supplementary figure 2). MIMIC-III and SafeICU (New Delhi) data served as external validation sets for adult and pediatric settings, respectively. MIMIC-III matched subset contains 22,247 numeric data files corresponding to 10,269 subject ids with vital time-series data. Subject-wise data is segregated for 17,294 ICU stays using ICU in-time and out-time information from the ICUSTAY table (Supplementary figure 1b). Inclusion and exclusion criteria and case-control cohort extraction criteria were the same as eICU (Supplementary figure 1b, 1c). The mean and standard deviation values of the physiological variables used in the modeling are listed in table1.

**Table1.**
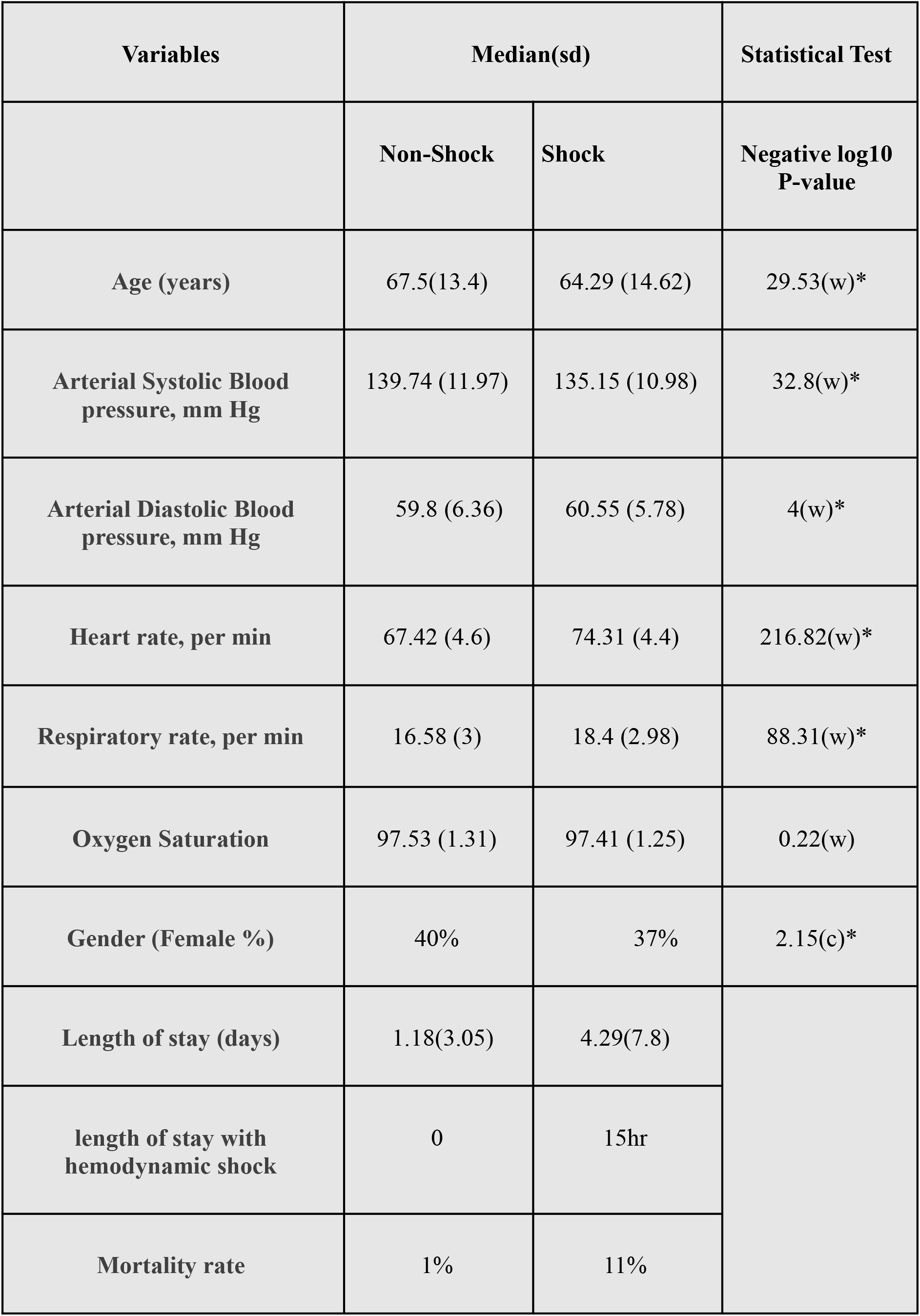
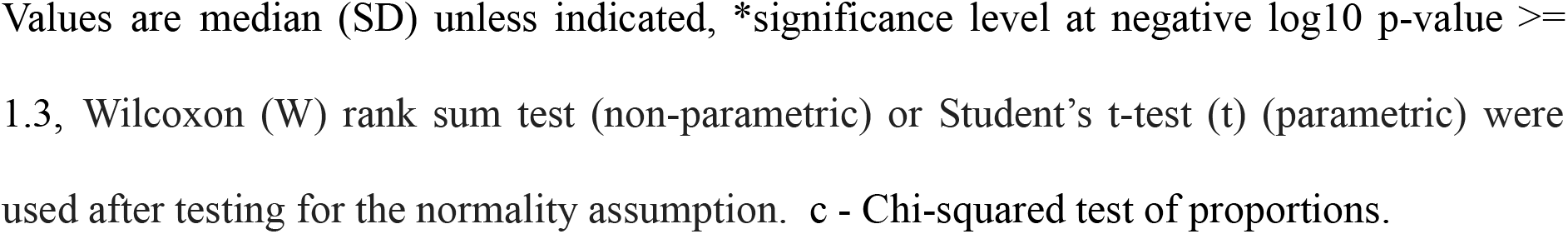
Cohort characteristics

| Variables | Median(sd) |  | Statistical Test |
| --- | --- | --- | --- |
|  | Non-Shock | Shock | Negative log10 P-value |
| Age (years) | 67.5(13.4) | 64.29 (14.62) | 29.53(w)* |
| Arterial Systolic Blood pressure, mm Hg | 139.74 (11.97) | 135.15 (10.98) | 32.8(w)* |
| Arterial Diastolic Blood pressure, mm Hg | 59.8 (6.36) | 60.55 (5.78) | 4(w)* |
| Heart rate, per min | 67.42 (4.6) | 74.31 (4.4) | 216.82(w)* |
| Respiratory rate, per min | 16.58 (3) | 18.4 (2.98) | 88.31(w)* |
| Oxygen Saturation | 97.53 (1.31) | 97.41 (1.25) | 0.22(w) |
| Gender (Female %) | 40% | 37% | 2.15(c)* |
| Length of stay (days) | 1.18(3.05) | 4.29(7.8) |  |
| length of stay with hemodynamic shock | 0 | 15hr |  |
| Mortality rate | 1% | 11% |  |

Values are median (SD) unless indicated, \*significance level at negative log<sub>10</sub> p-value ≥ 1.3, Wilcoxon (W) rank sum test (non-parametric) or Student's t-test (t) (parametric) were used after testing for the normality assumption. c - Chi-squared test of proportions.

### Model development and performance on eICU data

We trained and tested models on the eICU’s hospital ID-wise 5 fold data splits. We found the gradient boosting classifier model as the best performing model (Supplementary figure 2). We termed our final model hemodynamic shock early warning system (hsEWS), which generates prediction every 5 minutes and interpretable SHAP values for the observational time window. Depending upon the presence and absence of arterial blood pressure, we created two types of models, called hsEWS-invasive and hsEWS-non-invasive, respectively. The hsEWS-invasive model utilizes Arterial blood pressures (Systolic and Diastolic) achieved an AUROC of 86% and AUPRC of 93% (figure2 a. b.). hsEWS was found to identify 92% of all the hemodynamic shock events with an overall precision of 81% (figure2 b). The hsEWS-non-invasive model achieved an AUPRC of 90% and AUROC of 80%, and the overall recall was 90% with a precision of 79%. Therefore, for the subjects for whom arterial blood pressure is not being measured, a high recall hsEWS-non-invasive model can be applied. The model’s performance declined with increasing lead time; however, the recall remained above 89% for all lead times.

**Figure2:**
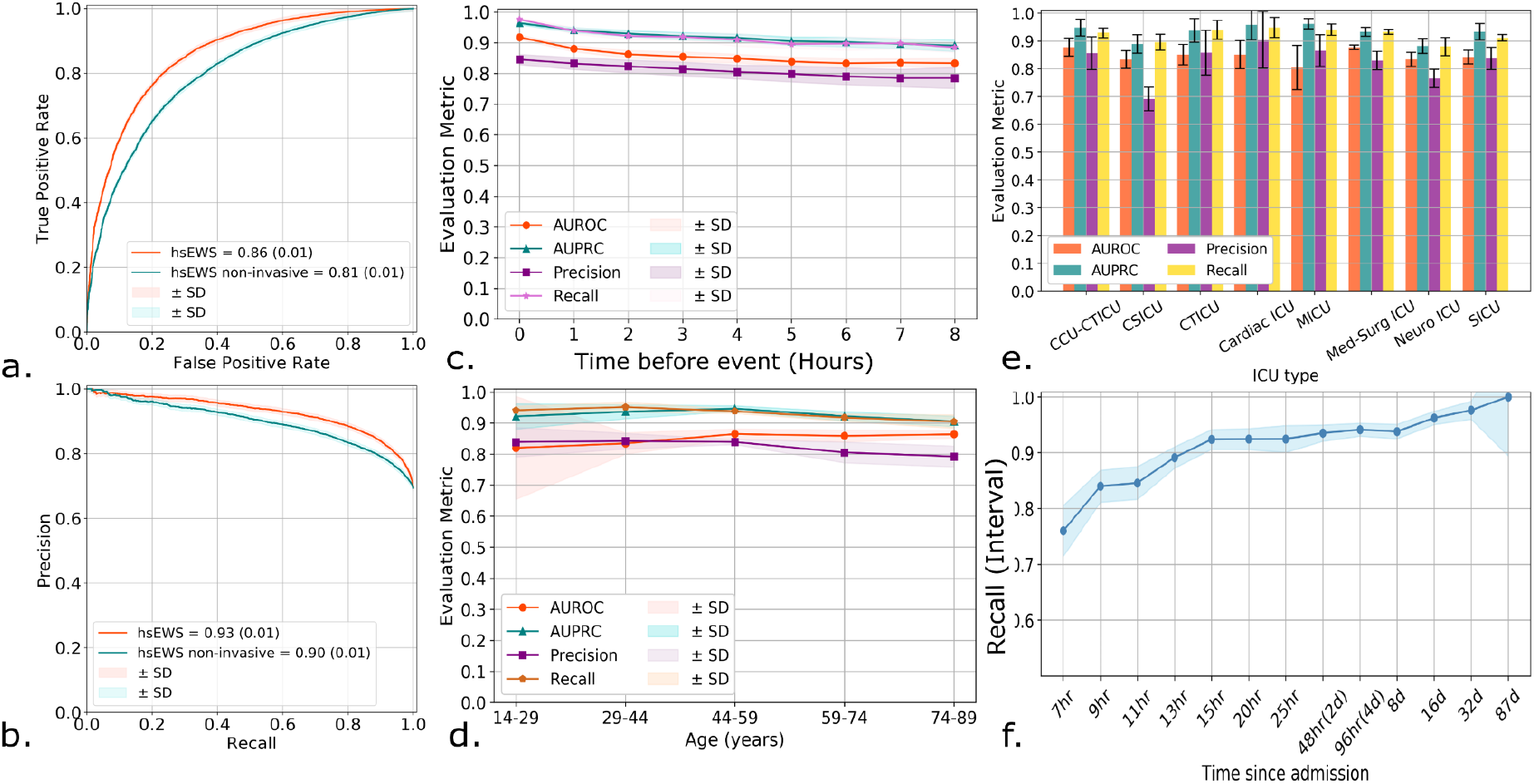
**a**.) AUROC for the prediction of Hemodynamic shock in the next 5 min to 8 hours. b.) AUPRC for the prediction of Hemodynamic shock in the next 5min to 8 hours. c.) Model performance for different lead times or times before the event. d.) Model performance for different age groups, eICU data contain adult age groups. e.) Model performance in different unit stay types (CCU – Coronary care unit, CSICU-Cardiac surgery ICU, CTICU-Cardiothoracic ICU, MICU-Medical ICU, SICU-Surgical ICU). f.) recall of the model for each time-interval shown on the x-axis for the prediction made during different lengths of stays in the ICU.

### Model performance for different age groups, unit types, and time-since admission

The model performance remained consistent for different age groups, with a slight decline in higher age groups above 60. However, AUROC increased for age groups above 60, and the model recall remained above 90%, with precision above 79% for all age groups (figure 2d.). We also evaluated performance for different types of ICU stays unit types. All the ICU types were found to have recall above 90% except for Cardiac Surgery ICU and Neurological ICU. The precision among these ICUs also declined below overall precision (figure 2e.). We also observed that recall of the model increases with length of stay, and the model started predicting the event with 90% recall after the 13th hour of stay.

### Feature Interpretation

A total of 3,970 hand-engineered time-series features were narrowed down to 2,120 important predictors using Boruta. This includes 51 classes of features listed in Supplementary Table S1. After developing the models, we computed SHAP values for interpreting the feature contribution on test predictions. We plotted the summary SHAP values of the top 15 contributors among all in figure 3a. The heart rate minimum and arterial systolic blood pressure minimum were found as the top predictors. The heart-rate minimum is positively correlated to its SHAP values (figure3.b). While the arterial systolic blood pressure minimum is negatively correlated to its SHAP values (figure 3c), we also found that aggregated linear trend was positively correlated to SHAP values, indicating an increased respiratory rate has an influence on the prediction of future hemodynamic shock. We also found features in the frequency domain using continuous wavelet transform (Cwt) coefficients, absolute energy, quintiles, mean, autocorrelation, a sum of recurring values, as important, complete list of selected features is presented in the Supplementary Table S1.

**Figure3.**
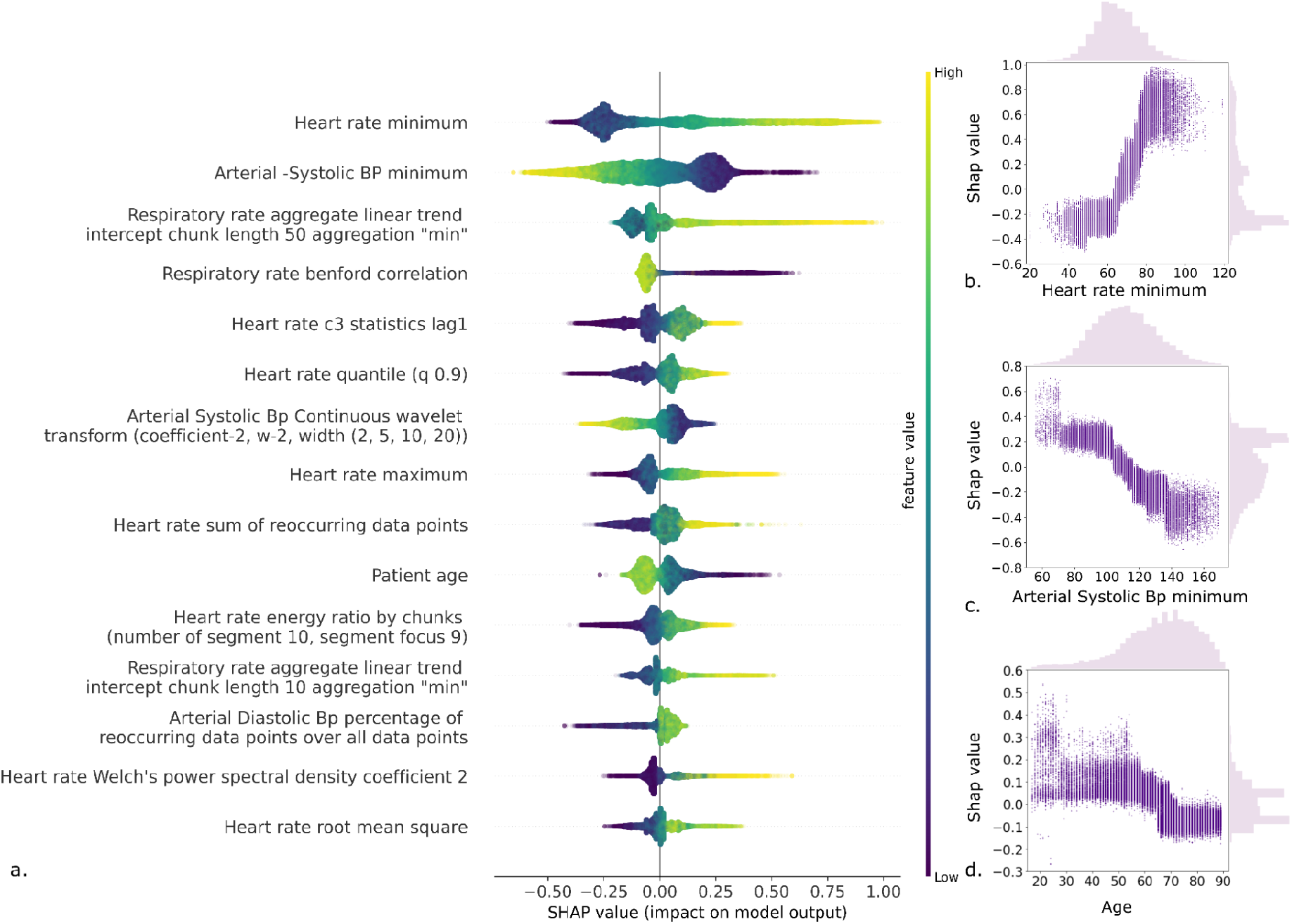
a.) SHAP values of top 15 hand-engineered time-series features (descending order) derived from the five-fold test set on eICU data. The Violin plot shows the distribution of SHAP values. The color map represents the feature value from low to high b.) A scatter plot of Heart rate minimum and corresponding SHAP values, showing a positive correlation between the two. c.) A scatter plot of Arterial Systolic Blood pressure minimum and corresponding SHAP values, showing negative correlation d.) A scatter plot of Patient age and corresponding SHAP values, showing a negative correlation.

### Performance of external validation of hsEWS model

We used two datasets, MIMIC-III (adult age groups) and SafeICU data (pediatric age group), for external validation. We found that with retraining on the new settings data, both in MIMIC and SafeICU, results improved over standalone performance and direct testing. With retraining hsEWS achieved an AUPRC of 93% on MIMIC (figure 4.a) and 91% on SafeICU (figure 4.b) and an AUROC of 87% both on MIMC and SafeICU (figure4 c, d). With direct testing (without retraining), hsEWS achieved an AUC of 78% and AUROC of 89% on MIMIC data. However, it performed poorly for direct testing on SafeICU pediatric data (figure 3b, 3d). As criteria for the pediatric hemodynamic shock are based on the age-specific cutoff, which differs from the adult population criteria, which could be one of the reasons for the poor performance. However, retraining of hsEWS on SafeICU pediatric training data resulted in an AUROC of 87% and AUPRC of 91% on a fivefold test-set (figure 3d). Overall, model recall remained consistent for all the lead-time, except for retraining on MIMIC, in which a decline in the recall is seen with lead-time (figure 4e). However, the retraining improved the precision on both MIMIC and SafeICU data cohorts by 10-15%, as shown in figure 4f.

**Figure 4.**
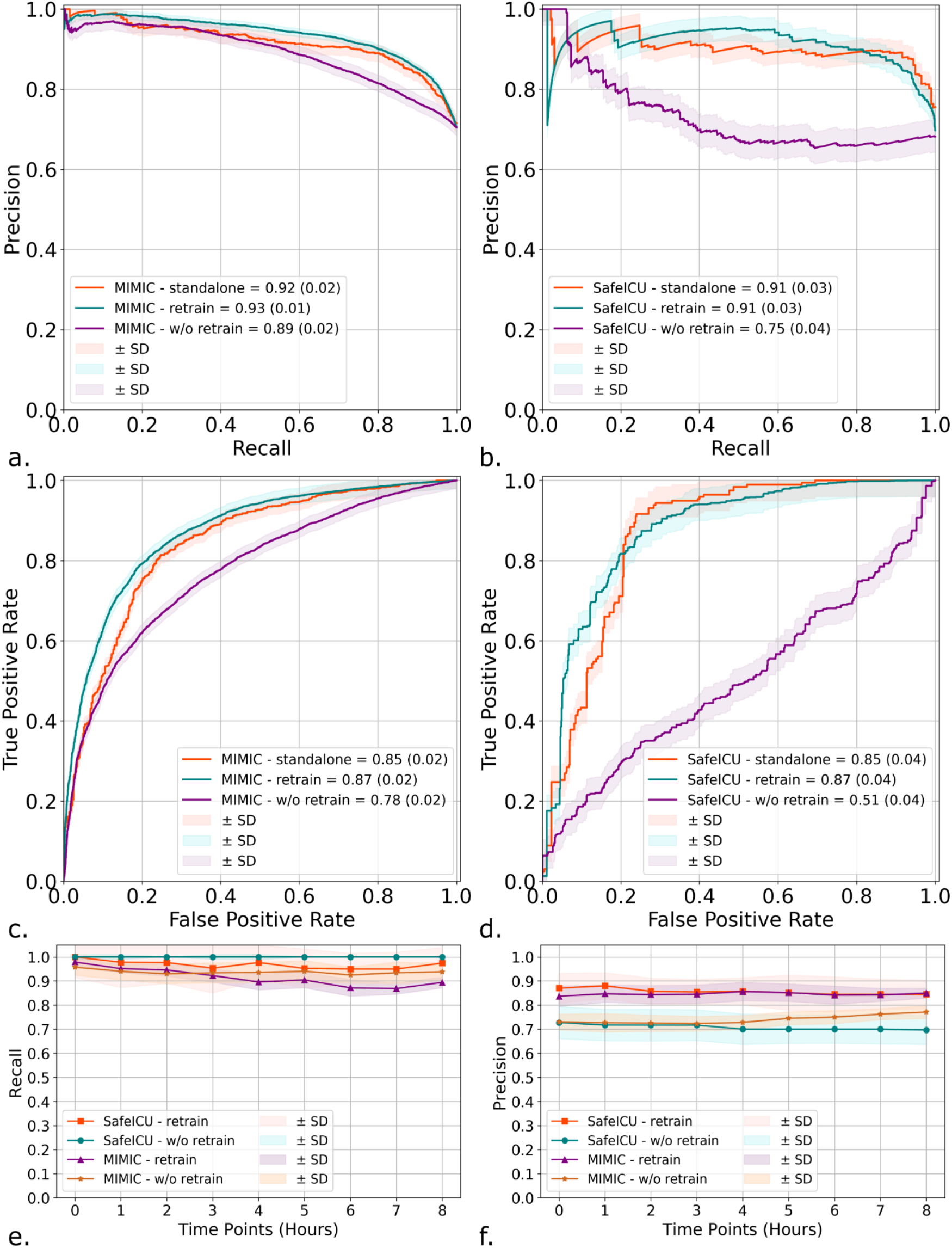
a.) AUPRC on MIMIC data for standalone, retraining, and without retraining (w/o retrain) of hsEWS model. b.) AUPRC on SafeICU data for standalone, retraining, and without retraining (w/o retrain) of hsEWS model. c.) AUROC on MIMIC data for standalone, retraining, and without retraining (w/o retrain) of hsEWS model. d.) AUPRC on SafeICU data for standalone, retraining, and without retraining (w/o retrain) of hsEWS model e.) Recall of retrained and without retrained hsEWS model on SafeICU and MIMIC data. e.) Precision of retrained and without retrained hsEWS model on SafeICU and MIMIC data.

### Performance of hsEWS retrained Model on different age groups, new setting’s ICU types, and time since admission

hsEWS retrained on MIMIC is tested on MIMIC’s test sets, and hsEWS retrained on SafeICU is tested on SafeICU test-sets. Models showed a consistent recall and precision above 90% and 81% respectively, on all age groups except for the 30-40 age group.. In the MIMIC cohort, performance with time since admission reached an interval recall above 90% from the 9th hour of stay, while in SafeICU, it remained consistently high with a recall of 100% from the beginning.

## Discussion

The development of machine learning based early warning systems for ICUs is not new^15,16,17,18^. However, most of these models have not made their way to successful deployments in the ICUs due to generalizability, interpretability, and workflow integration issues^25 26^. Further, it is generally expected that deep learning will outperform handcrafted features given sufficiently large data. Our research addresses these two challenges and shows for the first time that hemodynamic shock can be predicted in ICUs with models trained on patients with entirely different demographic, socio-cultural, and geographic factors. Further, our research opens the way to building interpretable models using handcrafted features which outperformed state-of-the-art deep learning models in predicting shock.

Clinically, the correlation between increased mortality in late-onset shock is indicative of a global under-recognition of this major killer in the ICUs despite reports suggesting a higher odds of mortality for late-onset of shock^27, 28^. We also found that a 10% increase in the time fraction of ICU stays spent in hemodynamic shock led to an estimated 6% increase in mortality rate. Our model Recall was found to increase for the late onset of shock (Figure 2e, 5c, 5d), so our model can reduce the mortality rate associated with late-onset of hemodynamic shock.

**Figure5:**
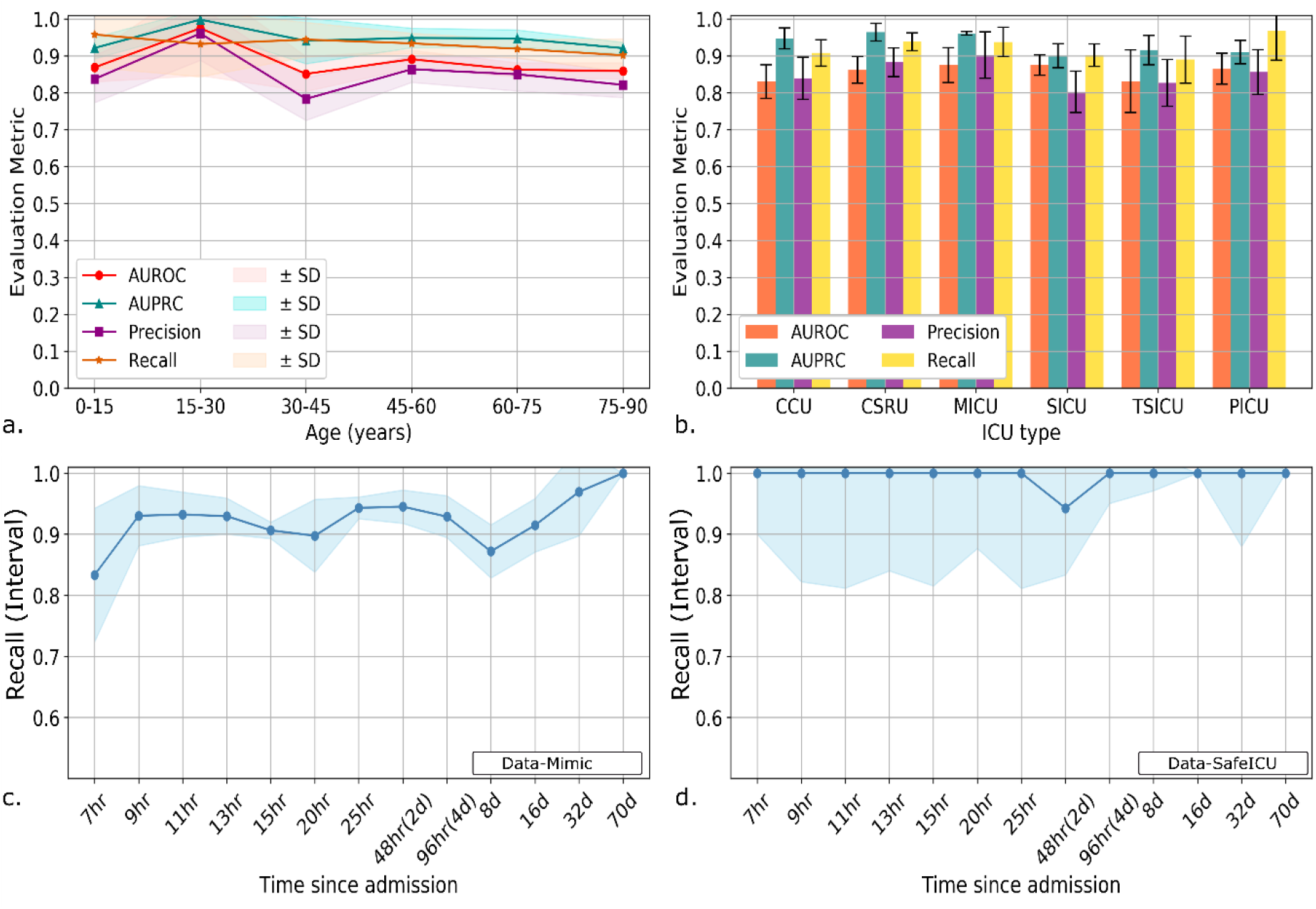
a.) Model performance parameters for different age groups (SafeICU -pediatric age group is embedded along with MIMIC-III (Adult age groups) data) b.) Performance of retrained hsEWS model in different first care units. c.) hsEWS retrained on MIMIC data, model Recall for “time since admission” d.) hsEWS retrained on SafeICU data, model Recall for “time since admission.”

In this work, we predicted hemodynamic shock using hand-engineered time-series features derived from 5 physiological time series. These time series were chosen due to their availability at the bedside across different age groups, geographies, and resource-limiting settings. It is often seen that many subjects are not monitored for arterial blood pressure. Therefore many models are not generalizable to those subjects. We, therefore, also trained a non-invasive hsEWS model which does not require invasive blood pressure measurements. Every hour of delays in detecting shock contributes to the increased risk of mortality in patients admitted to the ICU^6, 8^. Reliance on a large number of variables required to produce results hinders the model transfer for generalizability. In literature, a model with 36 variables to predict the onset of shock with an AUC of 82% has been previously reported^17^. Our model on pediatric ICU surpassed the performance reported in the literature by 5%, with a reduced number of variables required. These models have limitations of collecting many inputs that are not feasible for every minute resolution. We, in this work, used the readily available data to combat the issues related to large numbers of inputs. Our model uses the five vital signals data which is routinely monitored electronically. We have used the patient’s time series, which ensures to supply patterns in the patient’s physiology over a window of time, unlike a single time-point value. The hand-engineered time-series features were computed to represent the dynamics of the patient physiology described in Supplementary Table1.

We tested our model across age groups, ICU-types for different hospitals across the USA, and a pediatric ICU in India. Our assessment showed a consistent performance across these groups. We observed that the model performance decreases with an increase in the lead. However, our overall recall is 92%, at a prevalence of 83%.

In recent years, transfer learning has assumed increasing importance, especially in the low data regimes^29, 21, 30^. We transferred the hsEWS Model to the MIMIC-3 setting and SafeICU setting and show that transferring models trained on eICU data to MIMIC and SafeICU helped improve the overall AUC and precision while maintaining an overall higher recall above 90% (figure 5e and 5f).

Threshold computation was done to account for the population prevalence. Our model can be generalized by learning new population-based probability thresholds.

There are few limitations of our study. Our models are developed and tested on the retrospective data, and there is a need for testing the models prospectively. We have created an easy-to-use docker container that works well with real-time streaming data from monitoring. All the data were taken from a running ICU; therefore, almost all of the patients received some form of medication and might have come with a history of shock, which is not taken into account. However, this does not affect the prediction potential of our model since we made sure to include only subjects who developed shock in the unit. Also the training window or observational and lead-time data are free from any incidence of hemodynamic shock.

Finally, we believe that future directions should synergize hand-engineered and deep learning based predictions to improve patient outcomes. This will especially hold for multimodal data, including signals, text, tabular, and imaging data. While many handcrafted features in our work are straightforward to interpret, others may still need research for clinical interpretation. Here, we have offset this limitation by using Shapley Additive exPlanation (SHAP) value based model interpretation where we have computed SHAP values corresponding to each feature. These values can represent the influence of the feature on the prediction.

## Methods

### Dataset description & pre-processing

This study utilized two datasets available in the public domain and our pediatric data resource (SAFE-ICU). The eICU data^31^ is publicly available data containing more than 800 million observations for 1,92,751 patient admissions to 208 hospitals across the USA. It contains 12.2 million patient hours of vitals periodic data at the 5-minute resolution. Our inclusion criteria specified ICU stay >= 7.5 hours, absence of shock within the first seven hours of ICU admission, less than 10% missing data, and arterial blood pressure measurements. The second dataset, “Medical Information Mart for Intensive Care (MIMIC)” ^32^. MIMIC dataset hosted by physionet.org website as a publicly available data resource. Datasets are de-identified and available for analysis as per the MIT institutional review boards (IRBs) documented on the website. These data were collected between 2001 to 2012 at Beth Israel Deaconess Medical Center (BIDMC). We have used MIMIC III v1.4, which was released on September 2nd, 2016. Twenty-two thousand two hundred forty-seven numeric records that have been time-aligned and matched with 10,282 MIMIC III clinical database records were used for generating the Mimic Shock cohort. These data are summarized at 1-minute resolutions and merged using subject Ids. Further subject records were split into respective ICU stays based on the in-time and out-time given in the “ICUSTAYS” table of the clinical data of the MIMIC-III database.

The third dataset, SafeICU data resource^22^(Sepsis advanced forecasting engine ICU Database), is an in-house ICU data resource built at the Pediatric ICU of All India Institute of Medical Science, New Delhi. Study did not involve any contact or change in routine patient care. Hence a waiver of consent was sought and granted by the Institutional review board (IEC/NP-211/08.05.2015). SafeICU data collected between February 2016 and January 2020 were used to construct a final validation across the continent cohort.

### Imputation

All the time-series data were imputed using Kalman filter imputation^33^ up to 80% missingness. However, observation windows were selected in the final training based on the “not more than 10% imputation criteria”. The shock index was not imputed.

### Epoch and Cohort Generation

We created 30-minute epochs for dense labeling of the time interval as shock or no-shock. The labels for shock were derived using shock-index (SI). A time of onset of shock (t_(shock)_) is defined as the starting time of an epoch where the median shock index was greater than 0.7. We took time-series data of 420 minutes of the five signals; heart rate (HR), systolic arterial blood pressure (Sys-Abp), diastolic arterial blood pressure (Dia-Abp), respiratory rate, oxygen saturation (SpO2). A lead time of 0 to 8 hours at 1-hour intervals before the first occurrence of a 30-minute shock window was taken. Training data is further filtered based on 10% or less imputation. These scores can be computed for all timestamps present in the numeric data, thus precisely labeling the condition’s onset. For the pediatric ICU data, we labeled the data using an age-adjusted cutoff for shock index^34^.

### Model Development and Evaluation

eICU data cohort was used to train models to predict the next 0-8 hour shock status. Hospital IDs were randomly divided into five-fold cross-validation sets. Further training, validation, and test sets were chosen according to hospital ID. So every time, the model was tested on new 20% hospitals data. This was done in order to optimize models for generalization on external settings. Further, the Hand-engineered time-series features and raw signals data were transformed to z-scores for rescaling in order to facilitate model fitting. All the hyperparameters such as number of estimators, max features, epoch numbers, batch size, number of trees, and F1 threshold were optimized on the validation set. Final results were reported as the median and Standard deviation on five fold test sets.

### Hand-engineered times-series features extraction and selection

Five time-series signals; heart rate (HR), systolic arterial blood pressure (Sys-Abp), diastolic arterial blood pressure (Dia-Abp), respiratory rate, oxygen saturation (SpO2), were used to extract hand-engineered features. These features, also termed hand-engineered time-series (HETS) features, were extracted using the tsfresh python package^35^. This includes wavelet transform coefficients, Fourier transform coefficients, discriminative power, linear trends, recurrent value based features, etc. Python library “tsfresh” was used to calculate the features on the time-series data of the cohort. A total of 3970 features were extracted using the tsfresh python package. Variable selection was performed using Boruta, a feature selection algorithm, implemented through R package “Boruta” ^36^. Boruta selected features are listed in Supplementary Table S1.

### Modeling on hand-engineered time-series features

We built three models on hand-engineered time-series features, 1. Gradient boosting classifiers^37^, 2. Random-Forest models^38^ using sklearn (version 2.1.0) library in python 3.6. 3. Multilayer-perceptrons were trained on the hand-engineered time-series features selected after running feature selection algorithm Boruta. Hyperparameters tuning was performed using grid search for optimizing model performance parameters on the validation set.

### Modeling on raw signals

LSTM (long short-term memory) network is a type of recurrent neural network (RNN) that is capable of learning sequence information for temporal prediction problems^39^. As there will be lags of unknown duration between important events in a time series, LSTM is useful in time series data to process, classify and predict.

We tested two LSTM based architectures; first, CNN-LSTM contains convolutional neural networks followed by LSTM, connected to two densely connected layers of the perceptron to produce probability scores. Second, we used the Bi-Directional LSTM^40^ model. The model uses a time-distributed layer followed by a Bi-Directional LSTM layer which is further connected to three dense layers with tanh as activation function on the first two layers of dense and softmax at the last layer to get probabilities. The models were fitted in python 3.6 using Keras (version 2.3.1), Tensorflow (version 2.1.0). We used a single-layer perceptron for static features age and gender in parallel to the LSTM model. The weights of the last layer of the LSTM model and single-layer perceptron were concatenated. Finally, a multilayer perceptron with softmax activation was used to generate probability scores on the concatenated time-series and weighted static variable features.

### hsEWS and hsEWS-non-invasive model

The best model is selected from different classifiers mentioned above using AUROC and AUPRC. Selected Model was used for further evaluation; this model generates a score between 0 and 1 if found greater than a threshold attributed to future shock and non-shock risk. We computed this threshold on the validation set. The test is kept untouched. The model is termed as hsEWS (hemodynamic shock early warning system). hsEWS-non-invasive is another variant of the model in which invasive arterial blood pressures were not used.

### SHAP value computation for model interpretability

SHAP (SHapley Additive exPlanations) is a method that assigns each feature an importance value based on game-theoretic principles^41^. The SHAP values represent the influence of features on model prediction. We computed the SHAP value distribution of each feature for all the test sets. All the test sets were combined to plot the model explanation in terms of the top 15 important features and their relationship with SHAP values.

### Model generalization

We evaluated our models on two external validation sets. For this, we used MIMIC-III and SafeICU (Pediatric) data. We performed similar preprocessing with slight data-specific variation and cohort building as done for eICU. The higher resolution MIMIC Data and safeICU data were brought to a 5-minute resolution to match eICU resolution. We tested three scenarios of external validation—first, a standalone model, where the model is trained on the new setting’s data itself. Second, the model was retrained on the new setting’s training data and tested on the five-fold test sets. Third, we tested the hsEWS model directly on external validation data. For data specific prevalence-based probability threshold, we used the validation set of the external validation set. We choose the threshold over the precision-recall curve, which gives the maximum F1 score to optimize precision and recall.

## Data Availability

Data is available on reasonable request to the corresponding author.

## Acknowledgments

This work was supported by the Wellcome Trust/DBT India Alliance Fellowship IA/CPHE/14/1/501504 awarded to Tavpritesh Sethi. We thank Dr. Anurag Agrawal, Director, CSIR-Institute of Genomics and Integrative Biology for his valuable suggestions while preparing the manuscript. We also thank Mr. Varun Prakash and Mr. Anil Sharma for the technical support provided at PICU, AIIMS, and New Delhi.

The funders had no role in the execution of this study or the interpretation of the results.

## Author contributions

AN, TS designed the study. AN, PS, TS, RL were involved in acquiring data. AN, PS performed preprocessing and developed the cohorts, AN, PS, SF, AG involved in feature computation, feature selection AN, SF, PS, AG, AK, AS Developed models and performed model generalization, AN, VV formal analysis and visualized the results, AN, VV, RA involved in statistical analysis. WB, AMK performed multi-feature model experiments, AN, TS interpreted the data and wrote the first draft of the report. TS, AN, RL revised the report critically for important intellectual content. HB, PA, AN involved in the Docker App development.

## Conflicts of Interest and Source of Funding

The authors declare that they have no competing interests. This work was supported by the Wellcome Trust/DBT India Alliance Fellowship IA/CPHE/14/1/501504 awarded to Tavpritesh Sethi. Mr. Aditya Nagori is supported by CSIR-GATE fellowship. Mr. Pradeep Singh is supported through the Indo-Israel collaborative research grant received by Dr. Tavpritesh Sethi and Dr. Rakesh Lodha.

## Notes

### Competing Interest Statement

The authors have declared no competing interest.

### Author Declarations

"Datasets are de-identified and available for analysis as per the approval by MIT institutional review boards (IRBs) documented on the website http://physionet.org/. For SafeICU data ethical permission was granted by the All India Institute of Medical Sciences(AIIMS), New Delhi Institutional review board.

### Summary of Updates

Results improved, validation on new data and Language.

