## Supplementary Material for "Generalized Prediction of Hemodynamic Shock in Intensive Care Units"

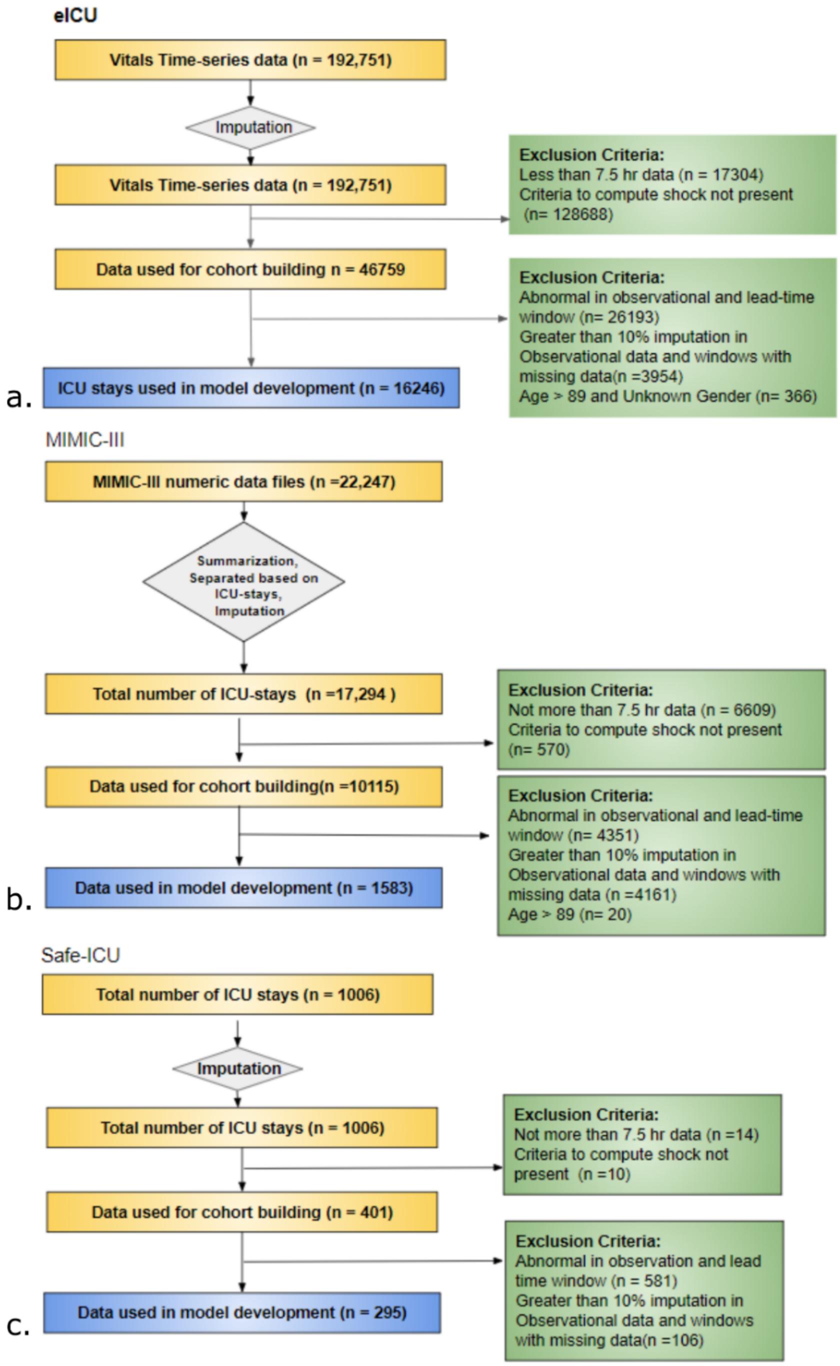


**Supplementary Figure S1**: Pre-processing of a.) eICU, b.) MIMIC-III and c.) SafeICU data for cohort building.

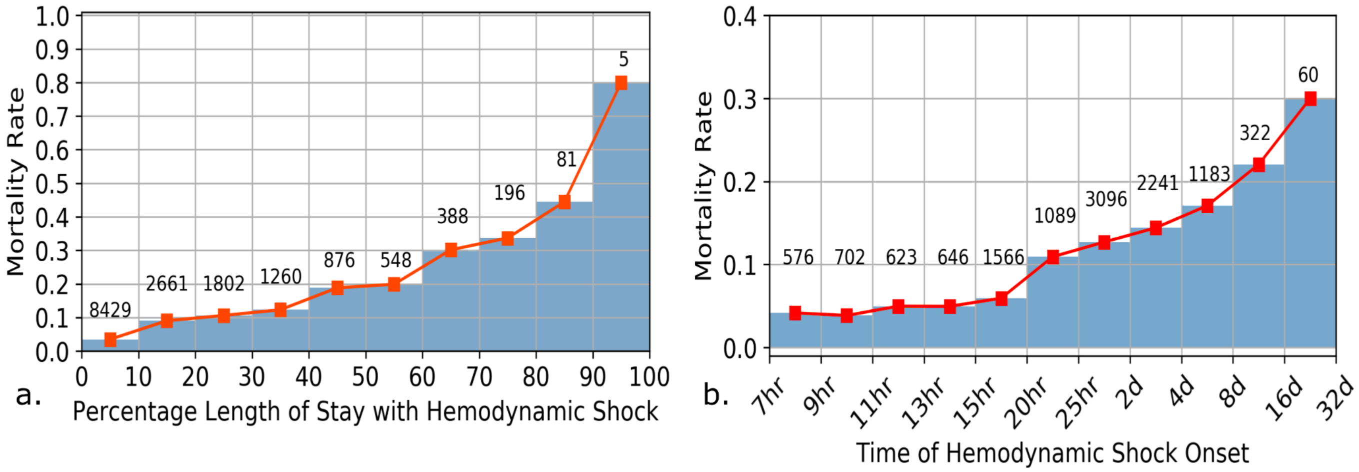


**Supplementary Figure 2**:. a.) Relationship between Mortality rate and Percentage of length of stay with hemodynamic shock.b.) Mortality rate vs time of onset of hemodynamic shock, it is observed that the mortality rate is higher in the stays in which onset happened later in ICU.

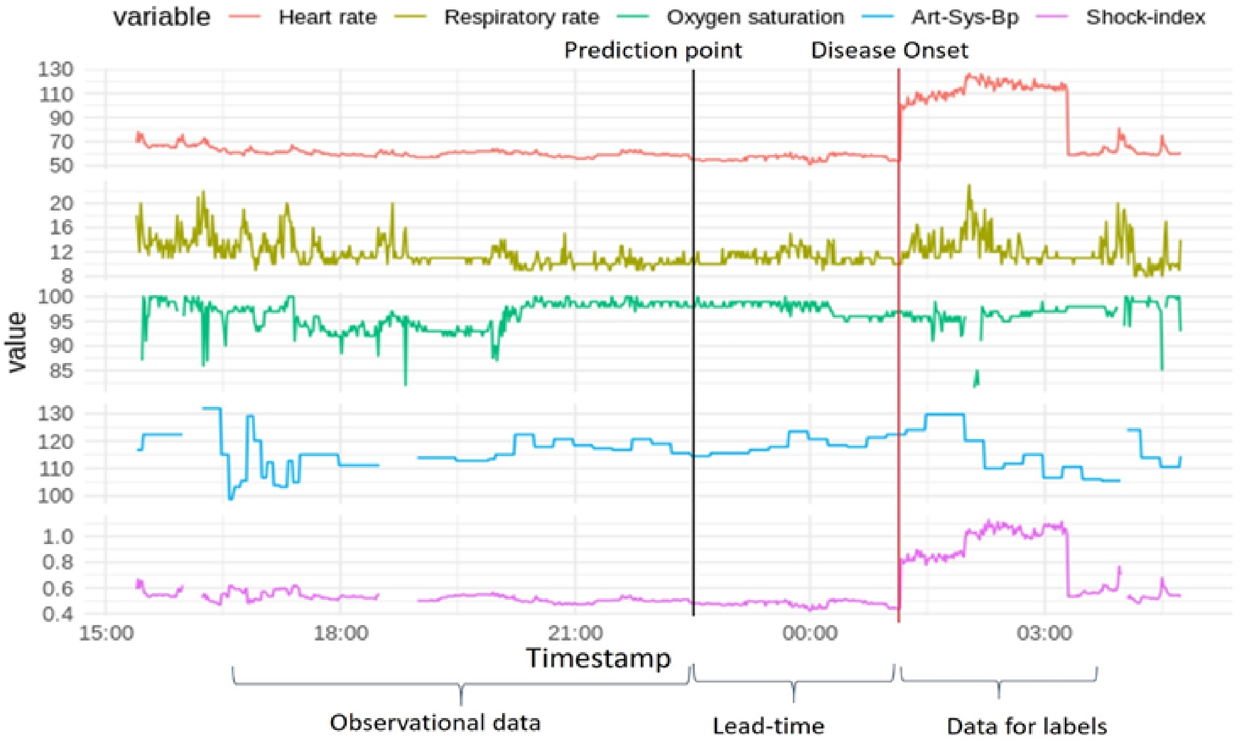


**Supplementary Figure 3:** Time-series data is labeled for each 30 minute time window as Shock positive or negative, a 420 minutes of observational length window is used to train models to predict the shock status for next 0-8 hours of lead time (Red line)

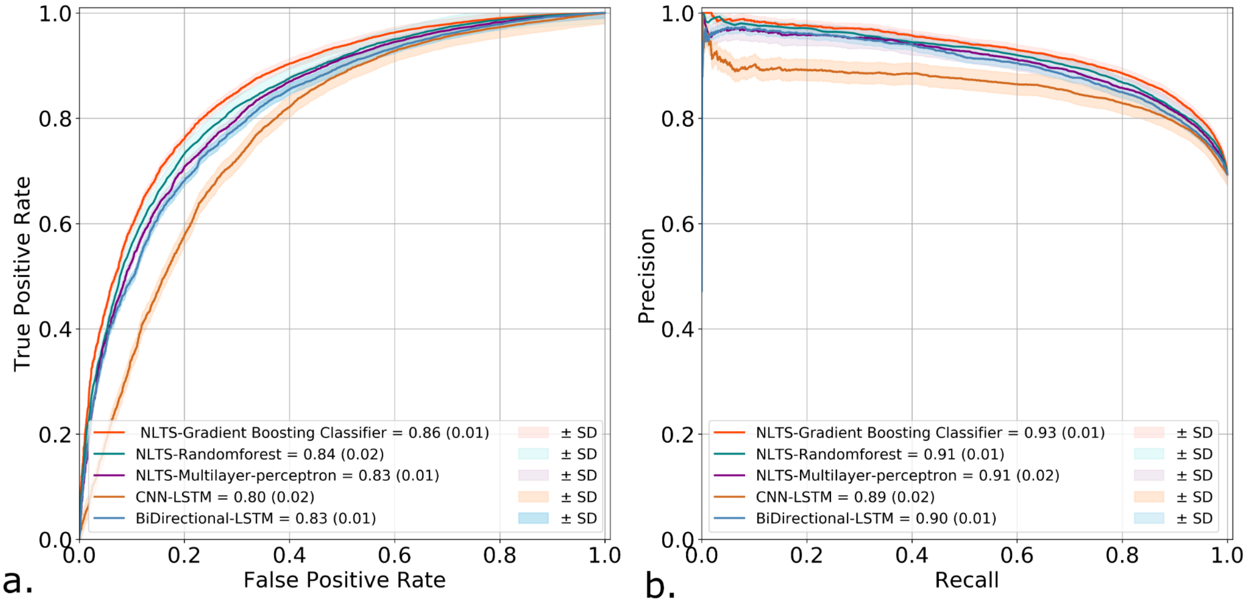


**Supplementary Figure S3:** a.) AUROC b.) AUPRC, different models trained on the eICU hospital splits. Gradient boosting classifier was found to be the best performing model.

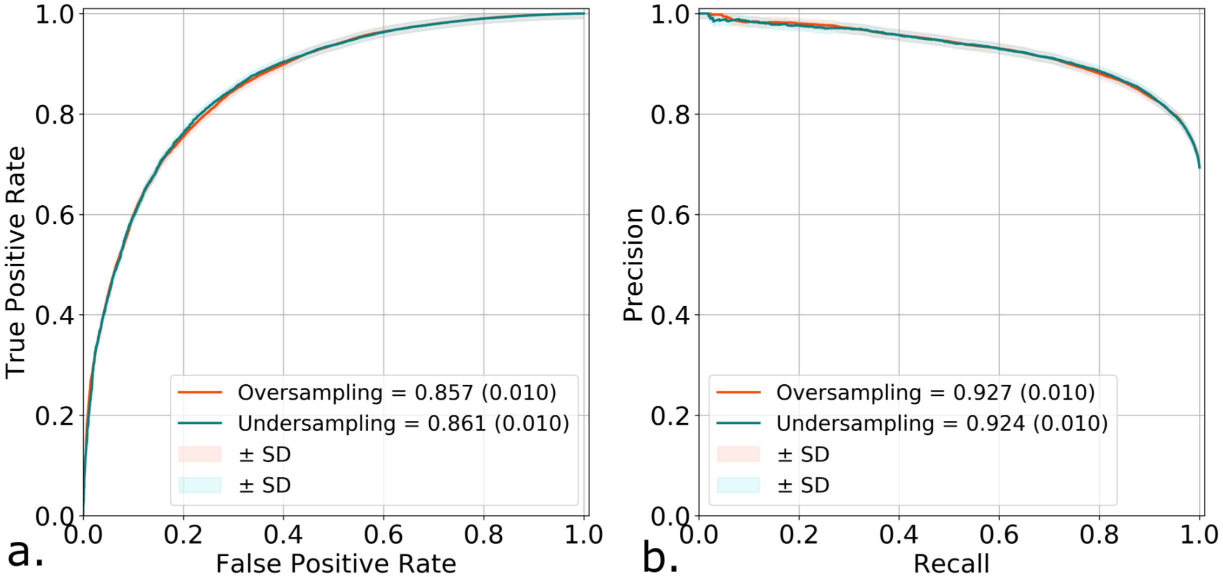


**Supplementary Figure4:** a.) AUROC b.) AUPRC, hsEWS model with over sampling of minority class, and random under sampling of the majority class.

**Supplementary Table S1:** Non-linear time-series features Description.

| SI.No. | Feature | Description |
|---|---|---|
| **1** | **Absolute Energy (abs_energy)** | Returns the absolute energy of the time series. |
| **2** | **Continuous Wavelet Transform Coefficients (cwt_coefficients)** | CWT proposes a time scale illustration of a signal. Duration of the examined signal will assist to dynamically identify non-linearities. |
| **3** | **Fast Fourier Transformation Coefficient (fft_coefficient)** | This feature uses the Fourier transformation algorithm to calculate the Fourier coefficients for the one-dimensional discrete Fourier transform. |
| **4** | **Mean** | This feature will return the mean of x. |
| **5** | **Quantile** | Calculates the q quantile of x. Where the quantile is dividing the sample into equal sized adjacent subgroups. |
| **6** | **Lag (c3)** | For all the time series perfect correlation will be at lag=0. Shift in the time series will decrease the correlation value. |
| **7** | **Sum of re-occurring data points (sum_of_reoccurring_datapoints)** | Returns the sum of all the time series data points that occur more than once. |
| **8** | **Sum (sum_values)** | Will calculate the sum over the time series values. |
| **9** | **Minimum** | Minimum number is the least value among the given set of values. |
| **10** | **Energy ratio by chunks (energy_ratio_by_chunks)** | This is expressed as a ratio of sum of all the squares of chunk i over the whole series of chunks. |
| **11** | **Partial Autocorrelation (Partial_autocorrelation)** | This provides the partial correlation for a time series in stationary condition. |
| **12** | **Aggregation linear trend (Agg_linear_trend)** | This calculates the value for an aggregation function of a linear least squares regression. |

| | | |
|---|---|---|
| **13** | **Linear trend (linear_trend)** | This will calculate the linear least-squares regression over one less than the length of the time series. |
| **14** | **Asymmetric statistic (time_reversal_asymmetry_statistic)** | It means that the entropy will increase as the time passes. |
| **15** | **Approximate entropy (approximate_entropy)** | It calculates the non-stationary and chaotic time series. |
| **16** | **Binned entropy (binned_entropy)** | It will first bin the values and then measures the entropy value. |
| **17** | **Maximum** | This feature calculates the highest among time series values. |
| **18** | **Mean change (mean_change)** | It provides the mean value of difference between time series values. |
| **19** | **Median** | This feature provides the median over time series values. |
| **20** | **Change quantiles (change_quantiles)** | First select the corridor on y axis and then within this corridor calculate the mean of the absolute change in series. |
| **21** | **Ratio beyond sigma (ratio_beyond_r_sigma)** | These are the ratio beyond the r sigma and are away from the mean of series. |
| **22** | **Index mass quantile (index_mass_quantile)** | These calculate the relative index where x% of mass of the time series is present at the left of the relative index. |
| **23** | **Sum of re-occurring values (sum_of_reoccurring_values)** | These are the sum of all the repeated values in the series. |
| **24** | **Longest strike below mean (longest_strike_below_mean)** | This is the length of the longest strike (consecutive subsequence) which is below the mean of the time series. |
| **25** | **Continuous Wavelet Transform peak (number_cwt_peaks)** | After the smoothing of time series with a ricker wavelet function, this feature provides the number of peaks in time series. |
| **26** | **Autocorrelation** | This provides the partial correlation for a time series in stationary condition. |
| **27** | **Largest fixed point of dynamics (max_langevin_fixed_point)** | When the time series is fitted to the Langevin model (with deterministic dynamics), this parameter gives the largest fixed point of the dynamics. |

| | | |
|---|---|---|
| **28** | **percentage_of_reoccurring_values_to_all_values** | In the given time series, this parameter gives a normalized ratio of all the unique values that are there in the time series more than one time. |
| **29** | **ratio_value_number_to_time_series_length** | This measure is closely related to percentage_of_reoccurring_values_to_all_values as this gives an output equal to 1 if every single value in the time series occurs only once. If the case isn't so, the output value would be : |
| **30** | **Standard_deviation** | Calculates the standard deviation of the time series. Expresses a measure which signifies how much all the values of the time series differ from the mean value of the time series. |
| **31** | **Variance** | Calculates the variance of the time series. It is the square of the standard deviation. |
| **32** | **Autoregressive coefficient (ar_coefficient)** | In a given time series, this first fits an autoregressive process's unconditional maximum likelihood to it, and then continues to calculate the AR coefficients. |
| **33** | **sample_entropy** | This parameter provides us with a measure of complexity. Closely related to approximate entropy, it has several advantages over it such as data length independence and lower computational cost. |
| **34** | **fft_aggregated** | This feature contains several of the properties of the time series such as skew, variance etc. |
| **35** | **kurtosis** | In a frequency distribution curve of a time series, this parameter gives a measure of how sharp the peak would be. |
| **36** | **Percentage_of_reoccurring_datapoints_to_all_datapoints** | In a given time series, this parameter gives a percentage of all the unique values that are there in the time series more than one time. This is different from percentage_of_reoccurring_values_to_all_values as in this percentage is normalized with respect to the unique values, rather than all of the data points. |
| **37** | **skewness** | Skewness gives a measure of the asymmetry in the time series, or in other terms shows the on which side does the time series lean towards (left or right). |
| **38** | **Cid_ce :** | This is the function calculator which estimates the complexity of time series |
| **39** | **Last_location_of_minimum :** | The location of the minimum value of x I'd calculated with respect to the length of x. |
| **40** | **Spkt_welch_density** | In this feature calculator, it first shifts the time series x from the time domain to the frequency domain. Then it estimates the cross-power spectral density |
| **41** | **Count_below_mean** | this function will return the number of values of x which may fall below the mean of x. |
| **42** | **agg_autocorrelation** | this function takes different lags into consideration and then calculates the value for an (fgg) aggregation function over the (R(l))autocorrelation |
| **43** | **Number_peaks :** | In time series x, this function will calculate the number of peaks which at the least support n |

| | | |
|---|---|---|
| **44** | **Absolute_sum_of_changes:** | This function returns the sum of the absolute value of the consecutive changes in the time series x |
| **45** | **Mean_abs_change** | This function returns the mean of the absolute difference between the subsequent time series value |
| **46** | **Longest_strike_above_mean:** | This function returns the length of the longest consecutive subsequence in x that which is bigger than the mean of x |
| **47** | **Friedrich_coefficients:** | It is the coefficient of the polynomial h(X),were h(x) is fitted to the deterministic dynamics of Langevin model |
| **48** | **Augmented_dickey_fuller:** | The test which checks if the unit root is present in a time series sample is called the augmented Dickey fuller test which is an hypothesis test. |
| **49** | **First_location_of_maximum:** | The first location of the maximum value of x is calculated with respect to the length of x. |
| **50** | **First_location_of_minimum** | The first location of the minimum value of x is calculated with respect to the length of x. |
| **51** | **Mean_second_derivative_central:** | this function will return the mean value for the central approximation of the second derivative |
